# Phase 2 dose-ranging study of the virologic efficacy and safety of the combination COVID-19 antibodies casirivimab and imdevimab in the outpatient setting

**DOI:** 10.1101/2021.11.09.21265912

**Authors:** Cynthia Portal-Celhay, Eduardo Forleo-Neto, Will Eagan, Bret J Musser, John D Davis, Kenneth C Turner, Thomas Norton, Andrea T Hooper, Jennifer D Hamilton, Cynthia Pan, Adnan Mahmood, Alina Baum, Christos A Kyratsous, Yunji Kim, Janie Parrino, Wendy Kampman, Lilia Roque-Guerrero, Roxana Stoici, Adil Fatakia, Yuhwen Soo, Gregory P Geba, Bari Kowal, A Thomas DiCioccio, Neil Stahl, Leah Lipsich, Ned Braunstein, Gary A Herman, George D Yancopoulos, David M Weinreich, Study Investigators

## Abstract

**Background:** The monoclonal antibody combination casirivimab and imdevimab (REGEN-COV^®^) reduced viral load, hospitalisation, or death when administered 1:1 as an intravenous (IV) dose ≥1200 mg in a phase 3 COVID-19 outpatient study. Availability of subcutaneous (SC) and/or lower IV doses should increase accessibility and/or drug supplies for patients.

**Methods:** This is a double-blind, placebo-controlled study of SARS-CoV-2-infected outpatients who were asymptomatic, or symptomatic but without risk factors for severe COVID-19. Patients were randomised to single IV dose (517 patients) of REGEN-COV 300, 600, 1200 or 2400 mg or placebo; or a single SC dose (286 patients) of REGEN-COV 600 or 1200 mg or placebo. The primary endpoint was time-weighted average daily change from baseline (TWACB) in viral load from day 1 (baseline) through day 7 in patients seronegative to SARS-CoV-2 at baseline.

**Findings:** All REGEN-COV treatments showed significant (p<0·001 versus pooled placebo) virologic reduction through day 7. Least-squares mean differences in TWACB viral load for the treatments versus placebo ranged from –0·56 to –0·71 log_10_ copies/mL. Each REGEN-COV treatment showed significant (p<0·001 versus pooled placebo) and similar virologic reduction through day 7. There were no safety concerns, dose-related safety findings, grade ≥2 infusion related/hypersensitivity reactions, grade ≥3 injection-site reactions, nor fatalities. Two serious adverse events not related to COVID-19 or the study drug were reported.

**Interpretation:** In asymptomatic and low-risk symptomatic SARS-CoV-2-infected outpatients seronegative for antibodies against SARS-CoV-2 at baseline, REGEN-COV significantly and comparably reduced viral load at all IV and SC doses.

**Funding:** Regeneron Pharmaceuticals, Inc. and Hoffman-La Roche

**RESEARCH IN CONTEXT:** *Evidence before this study:* Early phase 1/2 data in coronavirus disease 2019 (COVID-19) outpatients (NCT04425629) found that the REGEN-COV^®^ antibody combination, casirivimab and imdevimab, administered 1:1 as a single intravenous (IV) dose of 2400 mg or 8000 mg significantly reduced viral load over the first week compared to placebo. Enhanced viral clearance was more pronounced in patients who were seronegative for antibodies against severe acute respiratory syndrome coronavirus 2 (SARS-CoV-2), or who had high viral load at baseline. The phase 3 portion of this outpatient treatment study subsequently evaluated 1200 mg IV and 2400 mg IV doses, demonstrating consistent virologic efficacy, further demonstrating that REGEN-COV treatment reduced risk of COVID-19-related hospitalisation or all-cause death, and shortened time to symptom resolution. Virologic clearance was similar among those treated with any of the three doses (8000 mg, 2400 mg, or 1200 mg); therefore, maximal virologic efficacy may have been achieved at the 1200 mg dose in this treatment setting. These results warranted investigation of lower dose regimens.

*Added value of this study:* The present dose-ranging study evaluated whether a lower dose regimen could demonstrate virologic efficacy similar to that observed with 1200 mg IV and 2400 mg IV doses in outpatient treatment study. Exploration of a wider dose range will provide further characterisation of the clinical effects of REGEN-COV. Moreover, identifying a lower efficacious dose could bolster the ability to provide an adequate therapeutic supply of REGEN-COV in the setting of a global pandemic. A 1200 mg subcutaneous (SC) dose of REGEN-COV also prevented COVID-19 in household contacts of SARS-CoV-2-infected individuals (NCT04452318). The availability of a SC regimen could improve access for patients who have confirmed SARS-CoV-2 infection but for who IV infusion is not feasible.

*Implications of all the available evidence:* Despite the growing number of therapeutics with authorisation or approval for the treatment and/or prevention of COVID-19, there remains a significant global need for effective COVID-19 therapies. Additional therapeutics and dosing regimens will be required to meet demand and to meet the needs of specific patient populations. Lower IV doses of REGEN-COV, and the option of SC administration, should increase accessibility for patients. This increased availability needs to be weighed against several unanswered questions, including 1) whether the correlation between decreased viral load in the nasopharynx and improvement in clinical outcome holds at lower doses of REGEN-COV, and 2) whether the reduced drug exposure margins are sufficient to prevent viral escape and emergence of variants of concern.

## Introduction

Severe acute respiratory syndrome coronavirus 2 (SARS-CoV-2) first emerged in 2019 and is the causal agent of coronavirus disease 2019 (COVID-19), responsible for a world-wide pandemic.^1^ While some patients remain asymptomatic, others affected by COVID-19 are at risk of developing a range of respiratory conditions, from mild symptoms to severe and often fatal respiratory illness.^2,3^ Several factors, including age, pregnancy, race or ethnicity, and certain comorbidities (such as obesity, diabetes, cardiovascular disease, sickle cell disease, and chronic lung disease), place patients at high risk of more serious illness and increased hospitalisations.^4-6^ Recent studies among hospitalised patients found that high SARS-CoV-2 viral load is associated with increased mortality rates.^7,8^ Although similar studies are limited in the outpatient setting, such findings suggest that a SARS-CoV-2 antiviral therapy given to outpatients may reduce the risk of hospitalisation or death due to COVID-19.

All coronaviruses have spike (*S*) proteins that mediate entry into host cells by binding to angiotensin-converting enzyme 2 (ACE2) with high affinity.^9^ Blockade of host cell entry using neutralising antibodies against *S* proteins is one mechanistic strategy to reduce the pathogenesis of SARS-CoV-2.^10^ Casirivimab (REGN10933) and imdevimab (REGN10987) are human, immunoglobulin G1 monoclonal antibodies (mAbs) that simultaneously bind the receptor-binding domain of the SARS-CoV-2 *S* protein to block interaction with ACE2. In vitro and preclinical studies have shown that these two antibodies, administered together, minimise viral escape due to multiple SARS-CoV-2 mutations.^11-13^ These results point to a promising therapeutic approach of co-administering the antibodies 1:1 (REGEN-COV^®^ antibody combination) to reduce both the viral load of SARS-CoV-2 and COVID-19 disease progression.

In a phase 1/2 study outpatients positive for SARS-CoV-2 (NCT04425629), we previously reported that a single intravenous (IV) dose of REGEN-COV 2400 mg or 8000 mg reduced viral load, with a greater effect in patients who had not yet mounted an immune response (seronegative) or who had a high viral load at baseline.^14^ Based on these data, in November 2020, the 2400 mg IV dose of REGEN-COV received Emergency Use Authorization (EUA) from the United States Food and Drug Administration for the treatment of mild-to-moderate COVID-19 in adults and paediatric patients (aged 12–17 years, weighing ≥40 kg) who had positive SARS-CoV-2 viral testing and who were at high risk of progressing to severe COVID-19.^15^ At the same time that the trial described in this report was initiated, the phase 3 portion of the above outpatient treatment study (NCT04425629) using 1200 mg IV and 2400 mg IV doses confirmed the previous findings and further showed that REGEN-COV significantly reduced the risk COVID-19-related hospitalisation and all-cause death, as well as decreased the duration of symptoms. Therefore, in June 2021, based on data from the study presented in this report as well as the those presented above, the 1200 mg dose of REGEN-COV administered IV or SC received EUA for treatment of mild-to moderate COVID-19 and post-exposure prophylaxis in individuals who are at high risk for progression to severe COVID-19.^16^

A SC route of administration and lower doses could increase accessibility and supplies of drug. This current study evaluates the virologic efficacy and the safety of a range of IV and SC doses of REGEN-COV in outpatients with asymptomatic SARS-CoV-2 infection, or symptomatic with no risk factors for progressing to severe COVID-19.

## Methods

### Trial design

This phase 2, randomised, double-blind, placebo-controlled, parallel-group, dose-ranging study (NCT04666441) was conducted at 47 sites across the USA (**see appendix 1** for a full list of the study sites and investigators and **appendix 2** for the Regeneron study team).

Nasopharyngeal (NP) swabs and blood samples were collected from patients every other day for the first week. A phone visit occurred during the fourth week to collect safety information. After the first month, patients had monthly visits for 4 additional months. The final visit (end of study, day 169) was a phone call. Figure 1 depicts the schedule of events.

**Figure 1:**
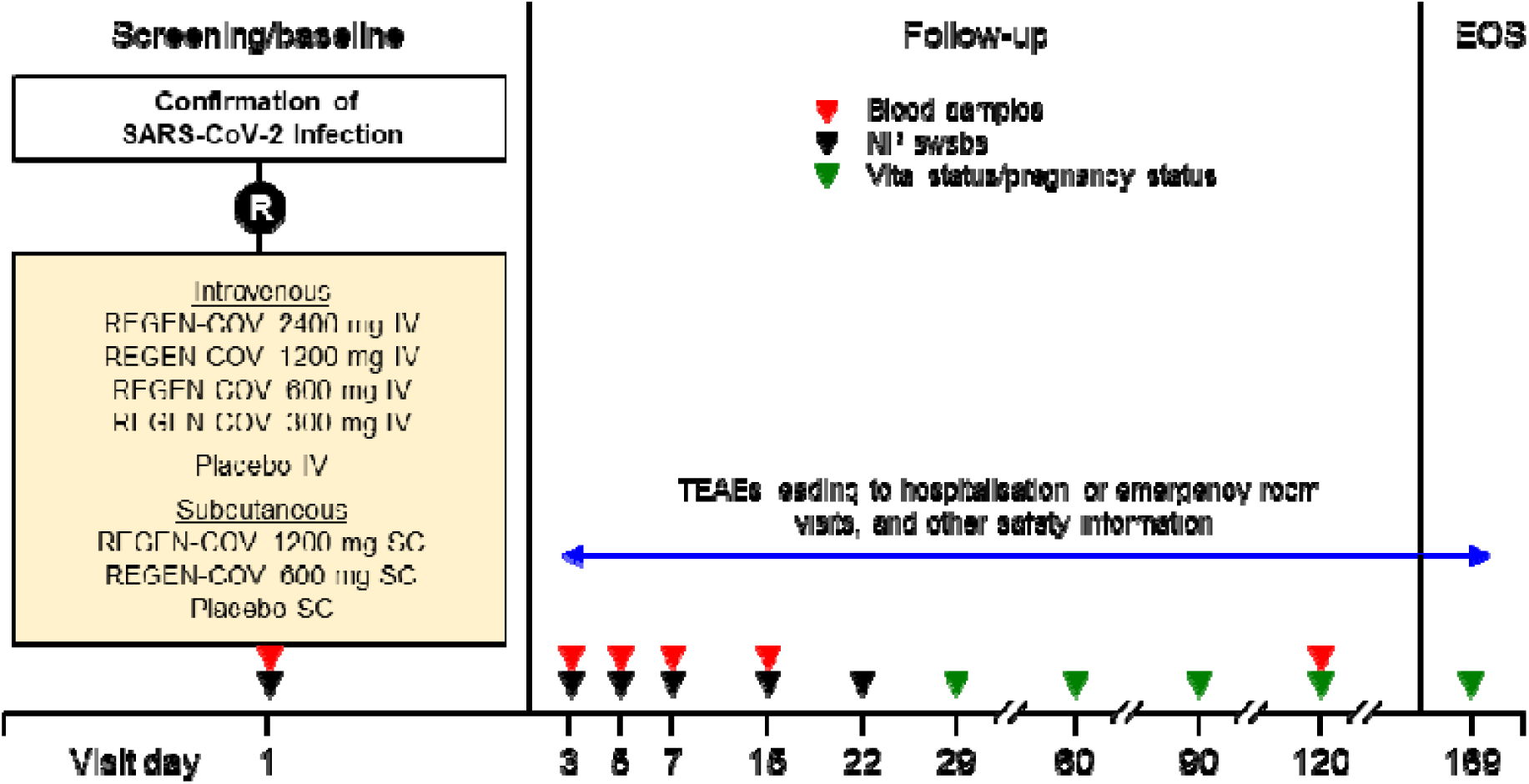
Study design. EOS=end of study; IV=intravenous; NP=nasopharyngeal; R=randomisation; SARS-CoV-2=severe acute respiratory syndrome coronavirus 2; SC=subcutaneous; TEAE=treatment-emergent adverse event.

### Patients

The study enrolled outpatients aged ≥18 years with a positive SARS-CoV-2 diagnostic test (using a local SARS-CoV-2 antigen, quantitative reverse transcription polymerase chain reaction [RT-qPCR], or other molecular diagnostic assay) from a sample (such as NP, nasal, oropharyngeal, or saliva) collected ≤72 h prior to randomisation. Eligible patients were SARS-CoV-2 infected but either asymptomatic or symptomatic ≤7 days before randomisation. Eligible symptomatic patients were required to be at low risk for developing severe COVID-19, defined as meeting all of the following criteria: body mass index <30 kg/m^2^; age ≤50 years; not pregnant; and no cardiovascular disease or hypertension, chronic lung disease or asthma, type 1 or type 2 diabetes mellitus, chronic kidney disease, or chronic liver disease. Eligible asymptomatic patients, with or without risk factors, could not have symptoms consistent with COVID-19 currently or at any time <2 months prior to randomisation. For the purposes of eligibility assessment, the presence or absence of COVID-19 symptoms was determined at the discretion of investigators. Immunosuppressed patents, as per the investigator’s assessment, were excluded from the study.

### Intervention and assessments

Eligible patients were randomised to receive a single dose of REGEN-COV or matching placebo administered by IV or SC route. Baseline (day 1) was the day of study drug or placebo administration. The IV single-dose regimens were REGEN-COV 300 mg (150 mg per mAb), 600 mg (300 mg per mAb), 1200 mg (600 mg per mAb) and 2400 mg (1200 mg per mAb). The SC single-dose regimens were REGEN-COV 600 mg (two injections of 300 mg per mAb) and REGEN-COV 1200 mg (four injections of 600 mg per mAb). Each injection volume was 2·5 mL. Healthcare providers, patients and their caregivers were blinded to placebo and the doses of REGEN-COV drug but not to the route of administration.

### Endpoints

The primary endpoint was the time-weighted average daily change from baseline (TWACB) in viral load (log_10_ copies/mL) from day 1 to day 7, as measured by RT-qPCR of NP swab samples, in patients who had a central-lab determined quantitative RT-PCR positive test at baseline and were seronegative (negative for anti-spike [S1] IgA, anti-spike [S1] IgG, and anti-nucleocapsid IgG) (**see appendix 3 and 4** for further details on methodology).

Secondary endpoints included the evaluation of additional indicators of virologic efficacy, safety, and tolerability, as well as REGEN-COV concentrations in serum over time (**see appendix 5** for pharmacokinetic analysis methods). The key safety variables included the incidence of all treatment-emergent adverse events (TEAEs) up to day 29, grade ≥3 TEAEs from day 30 and up to the end of the study (EOS), serious adverse events (SAEs) through EOS, and adverse events of special interest (AESIs). AESIs included all grade ≥2 infusion related reactions through day 4, grade ≥3 injection-site reactions through day 4, grade ≥2 hypersensitivity reactions through day 29, and any TEAE that led to a hospitalisation, or an emergency room visit throughout the study, regardless of relation to COVID-19.

### Statistical analysis

It was estimated that a sample size of 57 patients per treatment group would provide 98% power to detect a difference of –0·73 log_10_ copies/mL (assuming a standard deviation [SD] of 0·948) between any active treatment group and pooled placebo (IV and SC). The assumed SD and targeted difference used for powering was based on the observed variability and treatment difference of time-weighted average measurements in a companion phase 2 study of REGEN-COV in the outpatient treatment setting.^17^ With the assumption that 50% of enrolled patients would be seronegative at baseline, the study randomised approximately 800 patients to enrol 400 seronegative patients for efficacy analyses (ie, 57 patients per treatment group).

The route of administration was not expected to alter pharmacodynamic response in patients who received placebo. The IV and SC placebo groups were therefore pooled for all virologic efficacy analyses. Active treatment groups were not pooled for any efficacy analysis.

The overall modified full analysis set (mFAS) included all randomised patients who received treatment and who had a positive central-lab determined SARS-CoV-2 RT-qPCR result from NP swab samples at randomisation and was based on the treatment received. The primary analysis was conducted in the subset of patients in the overall mFAS who were seronegative at baseline for SARS-CoV-2 (seronegative mFAS). The efficacy analyses were based on observed data, with no imputation for missing data. The primary efficacy variable, TWACB, was calculated using the area under the curve (AUC) divided by the number of days (day 1 to day 7). AUC was calculated using the linear trapezoidal rule. Statistical analysis for viral load was conducted using a scale of log_10_ copies/mL. An analysis of covariance (ANCOVA) model with treatment group as a fixed effect, and baseline viral load and treatment by baseline interaction as covariates, was fitted to the data for analysis of the primary endpoint. The overall type I error was controlled at the 5% level via a pre-specified hierarchical testing procedure (ie, comparison between each active treatment group and placebo was formally tested). Additional statistical analysis details are described in the **appendix 6**.

The safety analysis set (SAF) included all randomised patients who received any study drug, and the pharmacokinetic analysis set included all patients who received any study drug and who had ≥1 non-missing drug concentration measurement following study drug administration.

### Role of the funding source

This study was supported by Regeneron Pharmaceuticals, Inc., and Hoffman-La Roche. Certain aspects of this project were funded in whole or in part with federal funds from the Department of Health and Human Services, Office of the Assistant Secretary for Preparedness and Response, Biomedical Advanced Research and Development Authority, under OT number: HHSO100201700020C.

### Study oversight

Study oversight is described in the **appendix 7**.

## Results

### Participants

A total of 815 patients were randomised and assigned to IV administration (n=523) or SC administration (n=292). Of those patients assigned to IV administration, 506 were SARS-CoV-2 positive via central-lab testing and therefore allocated to the overall mFAS; of those patients, 359 were seronegative at baseline for SARS-CoV-2 antibodies and therefore allocated to the seronegative mFAS. Of the patients assigned to SC administration, 304 were allocated to the overall mFAS, and 225 to the seronegative mFAS. In the analysis sets for efficacy (overall mFAS and seronegative mFAS), placebo IV and placebo SC are pooled, and this is reflected in the numbers presented above.

Of the seronegative mFAS patients assigned to IV administration, 80 were treated with 300 mg REGEN-COV, 68 were treated with 600 mg REGEN-COV, 72 were treated with 1200 mg REGEN-COV, 62 were treated with 2400 mg REGEN-COV, and 77 were treated with placebo (pooled placebo group) (table 1) (**see appendix, figure S1**). For the IV group, the mean (SD) age was 34·6 (9·6) years, 44·6% were male, 86·1% were White, 3·9% were Black, and 33·7% identified as Hispanic or Latino ethnicity. The mean (SD) baseline viral load was 7·2 (1·5) log_10_ copies/mL, and 56·8% of patients had a baseline viral load >10^7^ copies/mL.

**Table 1:**
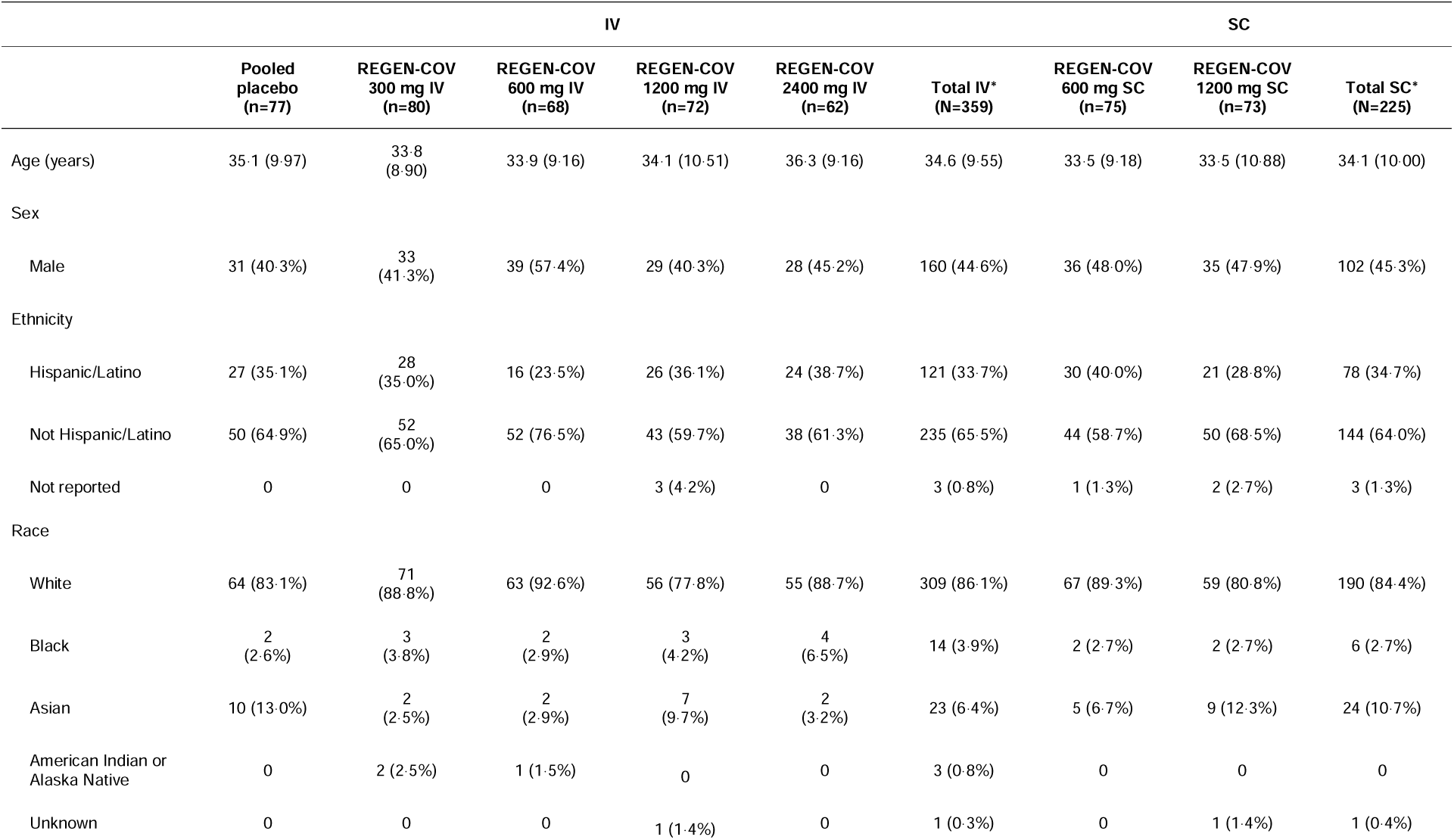
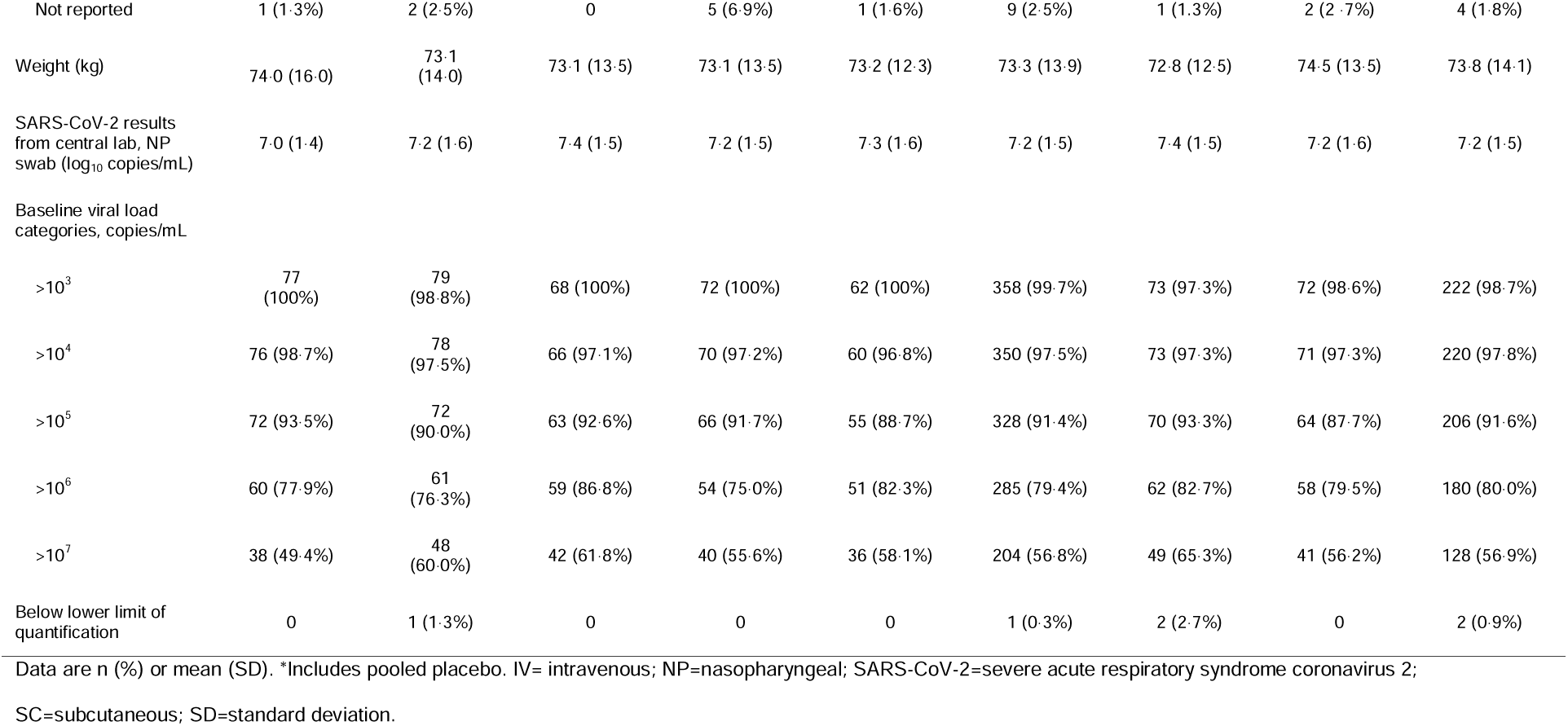
Demographics and baseline characteristics for seronegative mFAS IV patients and SC patients.

Of the seronegative mFAS patients assigned to SC administration, 75 were treated with 600 mg REGEN-COV, 73 were treated with 1200 mg REGEN-COV, and 77 were treated with placebo group (pooled placebo group) (table 1). For the total SC group, the mean (SD) age was 34·1 (10·0) years, 45·3% were male, 84·4% were White, 2·7% were Black, and 34·7% identified as Hispanic or Latino ethnicity. The mean (SD) baseline viral load was 7·2 (1·5) log_10_ copies/mL, and 56·9% of patients had a baseline viral load >10^7^ copies/mL.

Patient demographics and baseline characteristics were generally balanced between the IV and SC groups and pooled placebo (table 1). This was also true of the overall mFAS (n=712) and SAF (n=803; data not shown). Approximately 92·5% of the IV patients and 91·3% of the SC patients in the SAF were low-risk symptomatic patients, and the remaining were asymptomatic. All patients included in this analysis had the opportunity to complete the primary endpoint visit to day 7. A total of 195 IV patients and 106 SC patients completed visits up to day 29 (SAF) as of the data cut-off of February 8, 2021.

### Viral load

All REGEN-COV doses (SC and IV) significantly and similarly reduced viral load through day 7 versus pooled placebo (figure 2A). In the IV group, the least-squares mean (LSM) change in TWACB viral load (log_10_ copies/mL) between pooled placebo and REGEN-COV (95% CI) was –0·71 (–1·05, –0·38) (2400 mg), –0·56 (–0·89, –0·24) (1200 mg), –0·66 (–0·99, –0·34) (600 mg) and –0·57 (– 0·88, – 0·25) (300 mg). All differences versus placebo were statistically significant (figure 2A). In the SC group, the LSM change in viral load between pooled placebo and REGEN-COV (95% CI) was –0·56 (–0·87, –0·24) (1200 mg) and –0·56 (–0·88, –0·24) (600 mg). All differences versus pooled placebo were statistically significant (figure 2A). All pairwise comparisons between active treatment groups including different routes of administration demonstrated similar treatment effects (figure 2B).

**Figure 2:**
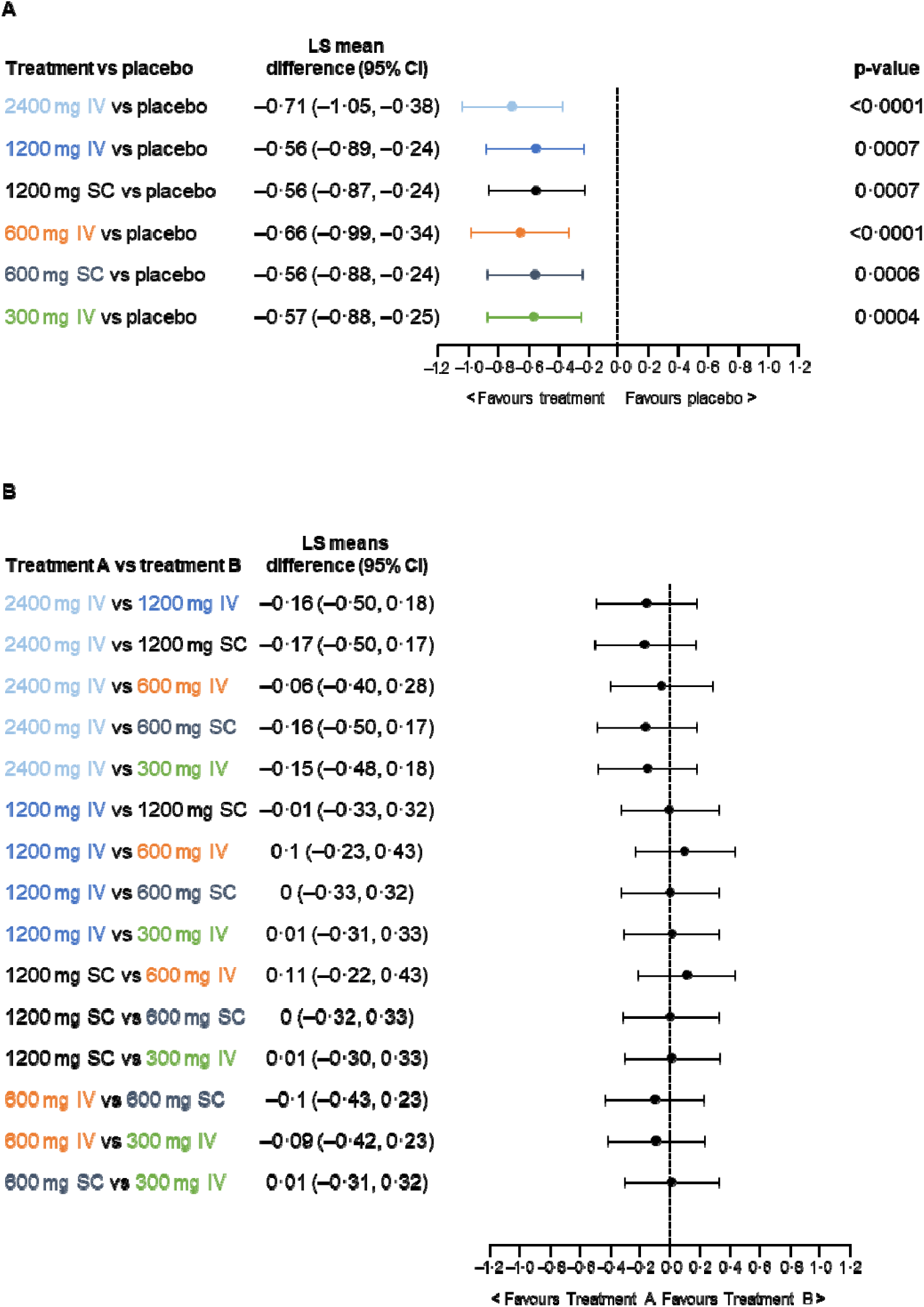
**Pairwise comparisons in the seronegative population of TWACB from day 1 to day 7 for each active treatment group versus (A) pooled placebo in the mFAS population or (B) another active treatment group in the PPS population** CI=confidence interval; IV=intravenous; LS=least squares; mFAS=modified full analysis set; PPS=per protocol set; SC=subcutaneous; TWACB=time-weighted average change from baseline.

All REGEN-COV doses (SC and IV) showed a similar reduction in viral load from baseline at each visit (figure 3). A difference in viral load between REGEN-COV treatments and pooled placebo was apparent by day 3, 48 h after administration of the study drug. In the seronegative mFAS, the LSM differences (log_10_ copies/mL) from pooled placebo on day 3 in the IV treated groups were –0·44 (300 mg), –0·74 (600 mg), –0·66 (1200 mg) and –0·53 (2400 mg). The corresponding differences in the SC treated groups were –0·34 (600 mg) and –0·49 (1200 mg).

**Figure 3:**
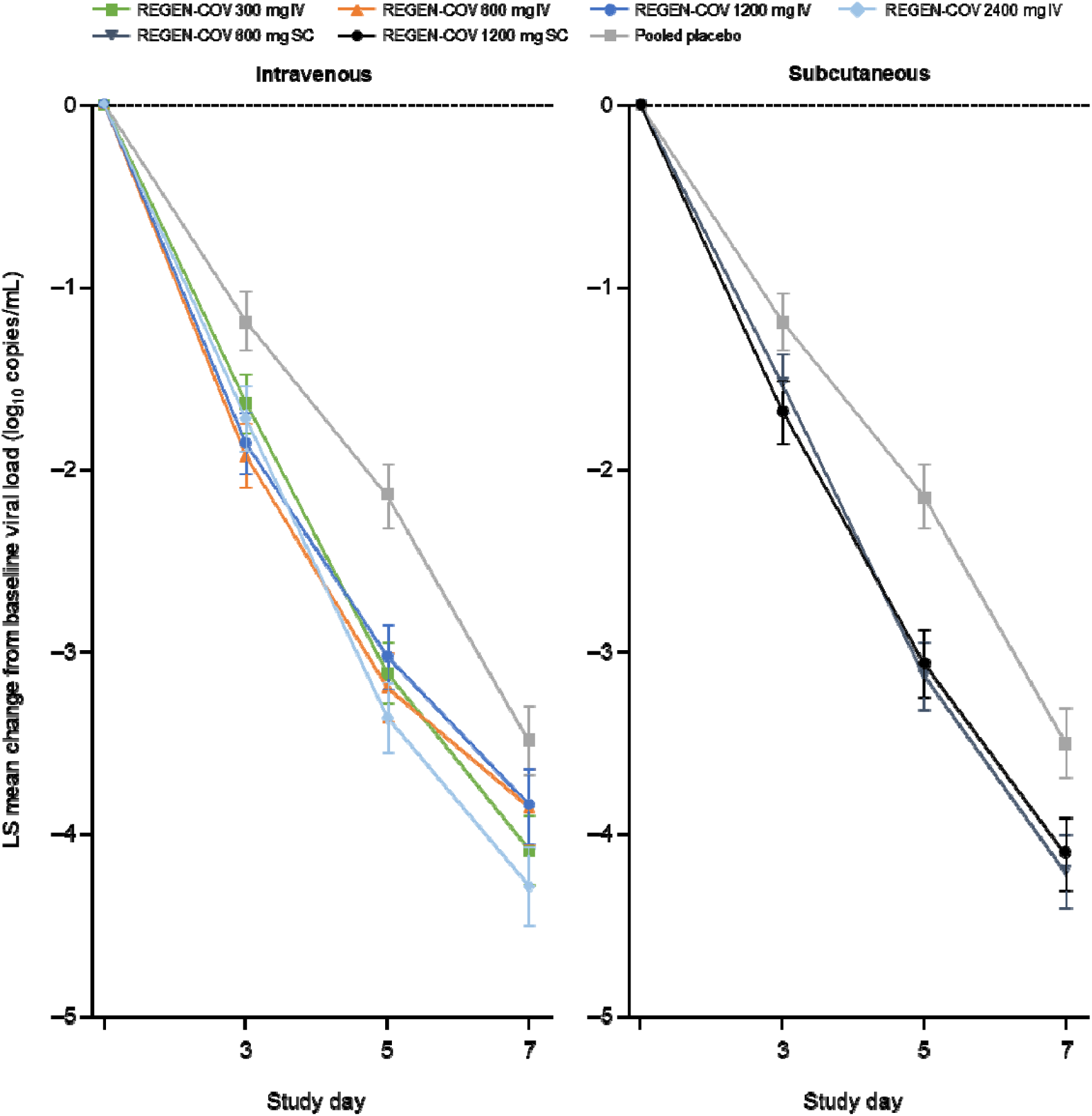
LS mean (± SE) change from baseline in viral load of IV and SC doses at each visit in the seronegative mFAS population. IV=intravenous; LSM=least-squares mean; mFAS=modified full analysis set; SC=subcutaneous; SE=standard error.

### Pharmacokinetics

The concentration–time profiles for each antibody were essentially superimposable from day 1 to day 7 for any given dose; the mean concentrations of REGEN-COV in serum are therefore presented as the sum of total casirivimab and total imdevimab concentrations. After a single IV dose, antibody concentrations fell monotonically from day 1 to day 7 (figure 4). For the SC doses, the mean concentrations of REGEN-COV reached near maximal values by day 3, with peak concentrations attained at day 7 (figure 4).

**Figure 4:**
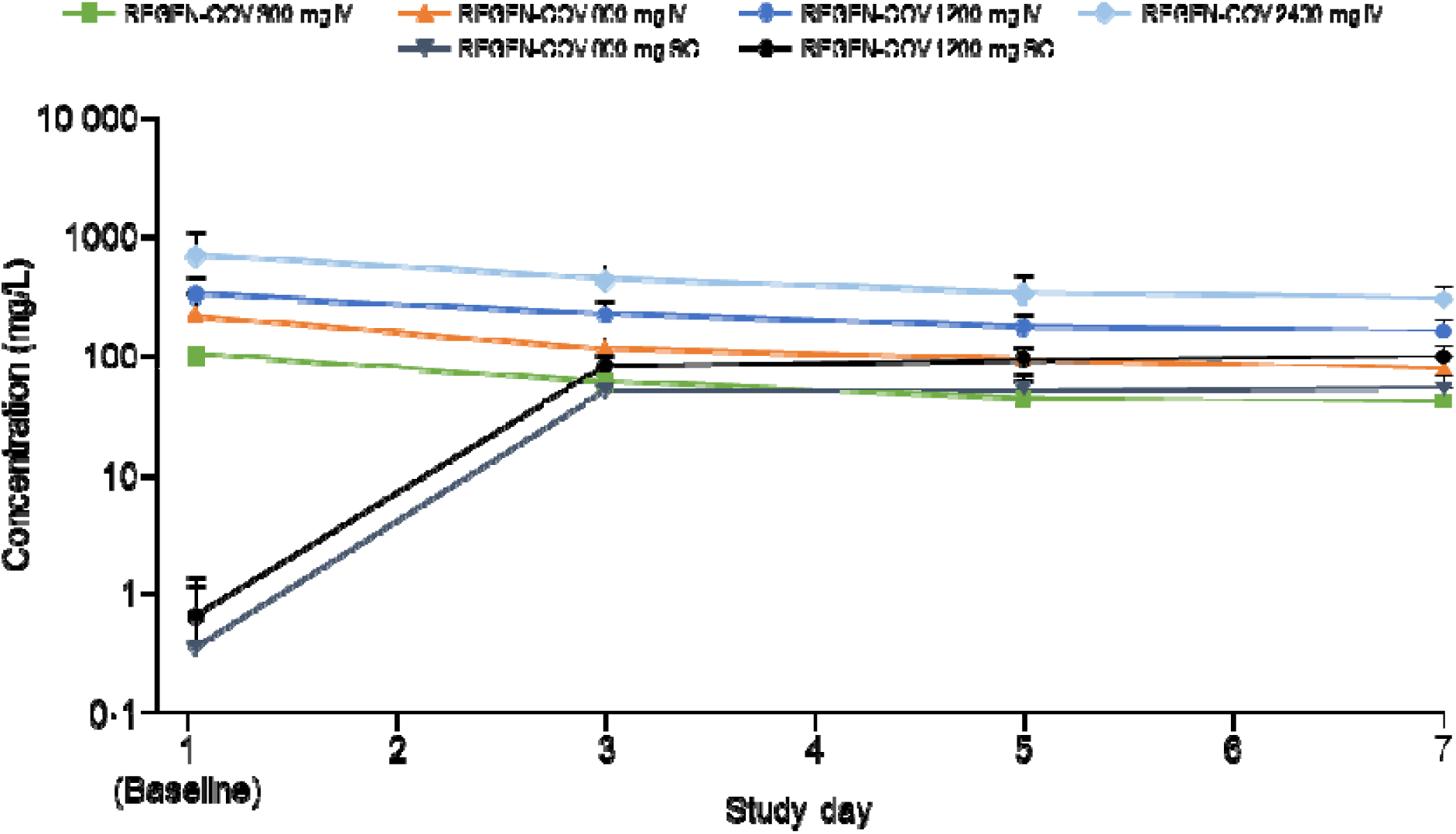
Mean (+SD) total REGEN-COV concentrations in serum in nominal time after single IV and SC doses. IV=intravenous; SC=subcutaneous; SD=standard deviation.

There was no association between TWACB in viral load versus total drug concentration through day 7 for the active IV or SC REGEN-COV doses, suggesting that maximum effect on viral load was achieved at all dose levels (figure 5).

**Figure 5:**
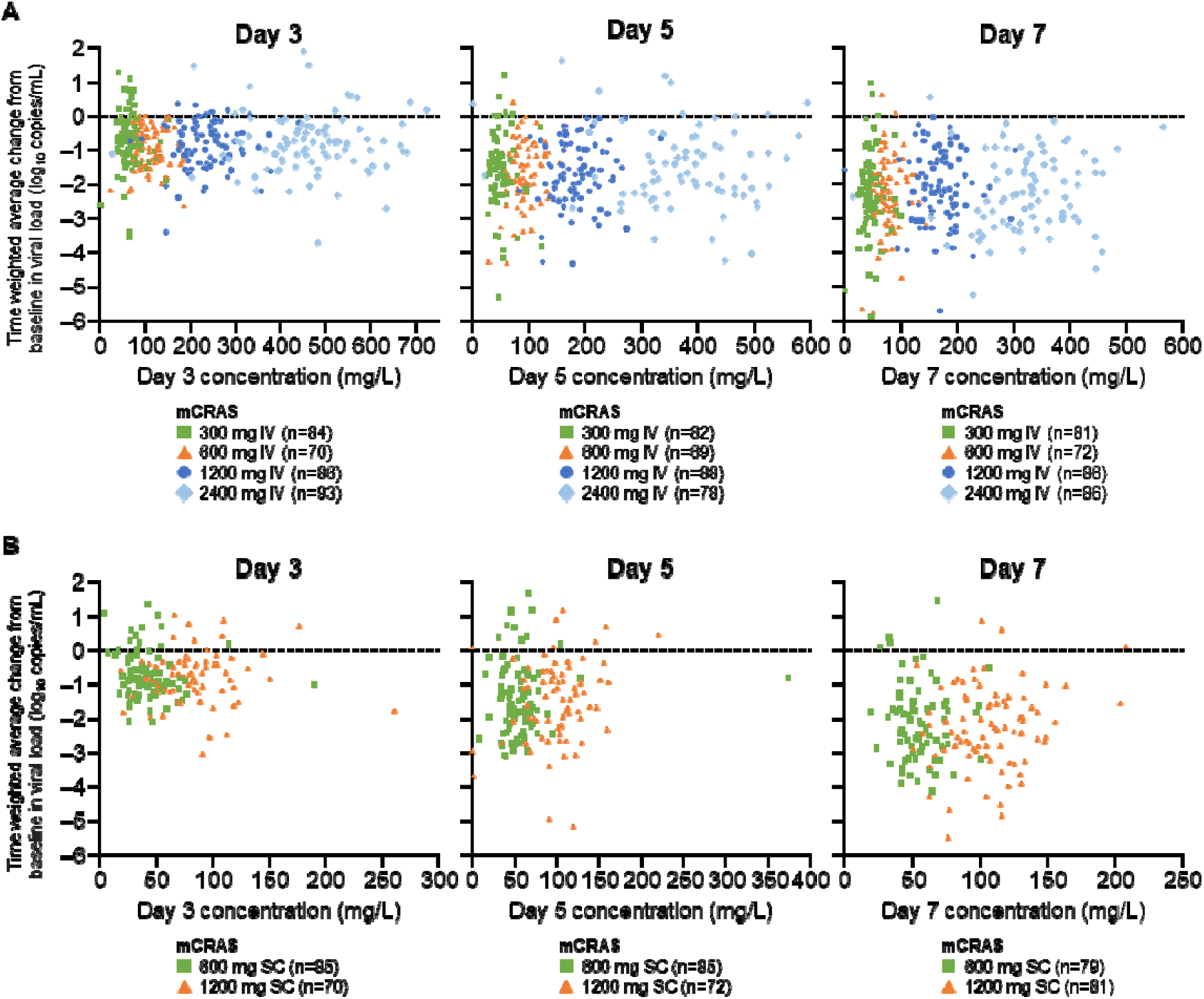
**TWACB in viral load (log_10_ copies/mL) versus total REGEN-COV concentrations* in serum in individual patients at days 3, 5, and 7 for (A) IV and (B) SC treatment groups (mCRAS)** *Total REGEN-COV concentration is the sum of total casirivimab and total imdevimab concentrations in serum. IV=intravenous; mCRAS=modified concentration-response analysis set; SC=subcutaneous; TWACB=time-weighted average change from baseline.

### Safety

Overall, in the safety analysis population (all randomised patients who received any study drug), all IV and SC REGEN-COV doses showed low rates of adverse events when compared with placebo. No serious safety concern was reported, and no dose-related safety findings were observed; no deaths occurred during the study (table 2). One patient did not complete infusion of the study drug, due to a TEAE of a mild, grade 1 infusion reaction from the 2400 mg IV dose, but the patient remained in the study. There were two SAEs reported in the 1200 mg IV and 2400 mg IV REGEN-COV groups, both of which were considered unrelated to the study drug or COVID-19. Both SAEs were miscarriages that occurred during the first trimester: one in primigravida and one in a patient with significant medical history of abortions. There were five additional pregnant women treated in study without any adverse outcome. No participant experienced a grade ≥2 infusion related reaction or hypersensitivity reaction, or grade ≥3 injection site reaction. The incidence of any TEAE that led to a hospitalisation or an emergency room visit (regardless of relation to COVID-19) was low, and none of the events were related to study treatment.

**Table 2:**
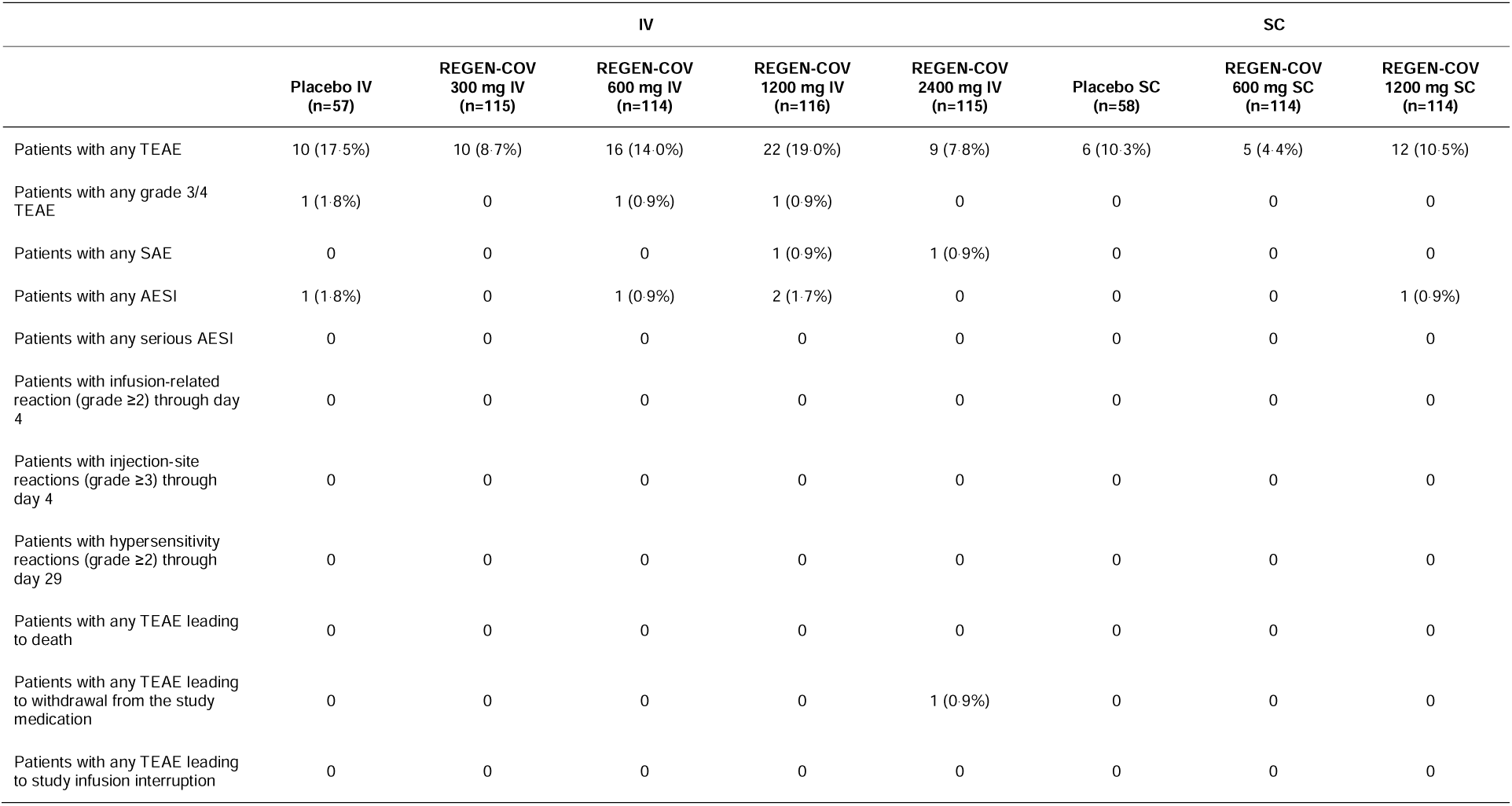
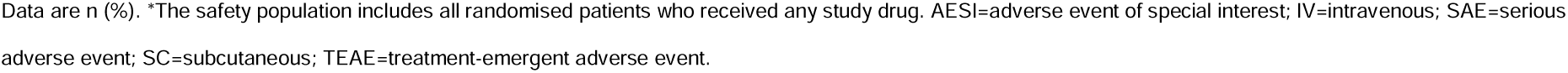
Adverse events in the safety population* in IV patients and SC patients.

|  | IV |  |  |  |  | SC |  |  |
| --- | --- | --- | --- | --- | --- | --- | --- | --- |
|  | Placebo IV<br>(n=57) | REGEN-COV<br>300 mg IV<br>(n=115) | REGEN-COV<br>600 mg IV<br>(n=114) | REGEN-COV<br>1200 mg IV<br>(n=116) | REGEN-COV<br>2400 mg IV<br>(n=115) | Placebo SC<br>(n=58) | REGEN-COV<br>600 mg SC<br>(n=114) | REGEN-COV<br>1200 mg SC<br>(n=114) |
| Patients with any TEAE | 10 (17.5%) | 10 (8.7%) | 16 (14.0%) | 22 (19.0%) | 9 (7.8%) | 6 (10.3%) | 5 (4.4%) | 12 (10.5%) |
| Patients with any grade 3/4 TEAE | 1 (1.8%) | 0 | 1 (0.9%) | 1 (0.9%) | 0 | 0 | 0 | 0 |
| Patients with any SAE | 0 | 0 | 0 | 1 (0.9%) | 1 (0.9%) | 0 | 0 | 0 |
| Patients with any AESI | 1 (1.8%) | 0 | 1 (0.9%) | 2 (1.7%) | 0 | 0 | 0 | 1 (0.9%) |
| Patients with any serious AESI | 0 | 0 | 0 | 0 | 0 | 0 | 0 | 0 |
| Patients with infusion-related reaction (grade ≥2) through day 4 | 0 | 0 | 0 | 0 | 0 | 0 | 0 | 0 |
| Patients with injection-site reactions (grade ≥3) through day 4 | 0 | 0 | 0 | 0 | 0 | 0 | 0 | 0 |
| Patients with hypersensitivity reactions (grade ≥2) through day 29 | 0 | 0 | 0 | 0 | 0 | 0 | 0 | 0 |
| Patients with any TEAE leading to death | 0 | 0 | 0 | 0 | 0 | 0 | 0 | 0 |
| Patients with any TEAE leading to withdrawal from the study medication | 0 | 0 | 0 | 0 | 1 (0.9%) | 0 | 0 | 0 |
| Patients with any TEAE leading to study infusion interruption | 0 | 0 | 0 | 0 | 0 | 0 | 0 | 0 |

Data are n (%). \*The safety population includes all randomised patients who received any study drug. AESI=adverse event of special interest; IV=intravenous; SAE=serious adverse event; SC=subcutaneous; TEAE=treatment-emergent adverse event.

## Discussion

In this phase 2 dose-ranging study of asymptomatic or low-risk symptomatic, outpatients who were SARS-CoV-2 RT-PCR positive and SARS-CoV-2 antibody seronegative at baseline, REGEN-COV treatment resulted in a significant reduction in viral load (day 1 to day 7) versus placebo. Viral load showed similar reductions across all REGEN-COV treatments evaluated, including the lowest doses of 300 mg (150 mg of each mAb) IV or 600 mg (300 mg of each mAb) SC. Reduction in viral load occurred as early as 48 h after the treatment administration (day 3 of measurement), decreasing (versus placebo) at all doses, including patients treated with the 600 mg SC dose. Although maximal concentrations in serum were attained a few days later for SC than IV dosing, REGEN-COV was rapidly absorbed into the systemic circulation following SC administration and achieved concentrations that provided comparable maximal virologic efficacy to the IV doses at the earliest time point measured.

Incidences of TEAEs, SAEs, and AESIs were low and balanced across the active drug and placebo groups, suggesting no difference in the REGEN-COV safety profile across all dose levels tested, whether delivered SC or IV. The pharmacokinetics of each antibody were linear and dose-proportional. Additional pharmacokinetic/pharmacodynamic modelling is underway to further evaluate the relationship of drug concentration with changes in viral load, in order to characterise the effect of REGN-COV on NP SARS-CoV-2 viral dynamics.

As mentioned earlier, we reported similar results on viral load and safety in a phase 1/2/3 outpatient clinical outcomes study of REGEN-COV administered as a 1200 mg, 2400 mg or 8000 mg single IV infusion (NCT04425629);^14^ the phase 3 data show equivalent clinical and virologic outcomes in the outpatient setting for 1200 mg and 2400 mg of REGEN-COV. Moreover, the 1200 mg SC dose demonstrated efficacy in a COVID-19 prevention and early treatment study of outpatients (NCT04452318).^18^ As a result of the totality of these data, Regeneron received an updated EUA for the treatment of mild-to-moderate COVID-19 and post-exposure prophylaxis in individuals who are at high risk of progressing to severe COVID-19 with a single 1200 mg IV or SC dose.^19^

The results of the present study suggest that the minimum dose for virologic efficacy may be lower than those evaluated. The current study did not determine whether the equivalent effect on reduction in viral load in the nasopharynx will correlate to similar improvements in clinical outcomes. In addition, while concentrations of each antibody on day 3 for the 600 mg SC dose are approximately 60 to several hundred times the concentrations required to neutralise wild-type SARS-CoV-2 virus and variants of concern *in vitro*, higher doses providing greater exposure margins are considered advantageous in the event that new variants emerge.

Despite the growing number of therapeutics with authorisation or approval for the treatment and/or prevention of COVID-19, there remains a significant global healthcare need for effective COVID-19 therapies. Multiple therapeutic and preventive agents are required to meet the demands and the needs of specific patient populations. Investigating different doses and methods of administration is key to characterising the dose-response for REGEN-COV and establishing a lower dose with similar effectiveness, while providing convenience for patients. Identifying lower doses that reduce viral load while not compromising clinical efficacy in preventing hospitalisation and death ensures the ability to provide therapeutics to as many individuals as possible.

In this analysis of low-risk patients (symptomatic with no significant risk factors, or asymptomatic) who were SARS-CoV-2 PCR-positive at baseline and seronegative, REGEN-COV significantly and comparably reduced viral load versus pooled placebo across all treatment groups, including at doses as low as 300 mg IV or 600 mg SC. REGEN-COV showed low levels of adverse events across all doses evaluated. Further studies are required to evaluate whether clinical outcomes remain the same with lower doses of REGEN-COV.

## Supporting information

Author COI

Author COI

Author COI

Author COI

Author COI

Author COI

Author COI

Author COI

Author COI

Author COI

Author COI

Author COI

Author COI

Author COI

Author COI

Author COI

Author COI

Author COI

Author COI

Author COI

Author COI

Author COI

Author COI

Author COI

Author COI

Author COI

Author COI

Author COI

Author COI

## Data Availability

Qualified researchers may request access to study documents (including the clinical study report, study protocol with any amendments, blank case report form and statistical analysis plan) that support the methods and findings reported in this manuscript. Individual anonymised participant data will be considered for sharing once the product and indication have been approved by major health authorities (eg, FDA, EMA, PMDA, etc.), if there is legal authority to share the data and there is not a reasonable likelihood of participant re-identification. Submit requests to https://vivli.org/.

## Contributors

CPC, EFN, WE, BJM, JDD, KCT, TN, ATH, JDH, AM, AB, CAK, YK, JP, WK, YS, GPG, BK, ATD, NS, LL, NB, GAH, GDY, and DMW contributed to study concept and design. CP, LRG, RS, AF were involved in data collection. CP provided administrative, technical or material support. WE, BJM, YS provided statistical analysis. CPC, EFN, WE, BJM, JDD, KCT, TN, ATH, JDH, AM, GPG, GAH, GDY, and DMW provided analysis and interpretation of the data. CPC, EFN, WE, KCT drafted the manuscript. All authors provided critical revision of the manuscript for important intellectual content and provided approval to submit.

## Declaration of interests

CPC, EFN, WE, BJM, JDD, TN, CP, AM, YK, JP, WK, YS, GPG, BK, ATD, LL, NB, and DMW are Regeneron employees/stockholders. KCT, JDH, and GAH are Regeneron employees/stockholders and have a patent pending, which has been licensed and receiving royalties, with Regeneron. AB, CAK, NS, and GDY are Regeneron employees/stockholders and have issued patents (U.S. Patent Nos. 10,787,501, 10,954,289, and 10,975,139) and pending patents, which have been licensed and receiving royalties, with Regeneron. ATH is a Regeneron employee/stockholder, former Pfizer employee and current stockholder, and has a patent pending, which has been licensed and receiving royalties, with Regeneron. LRG, RS, and AF have no conflicts to declare.

## Acknowledgements

The authors would like to thank the patients and their families, as well as all other study investigators and all investigational site members involved in this study. Caryn Trbovic, PhD, S. Balachandra Dass, PhD, and Brian Head, PhD of Regeneron Pharmaceuticals Inc., and Michele Bipath, PharmD, of Prime Global, assisted with the development of the manuscript. Helen Kang, PhD, and Travis Bernardo, PhD, of Regeneron Pharmaceuticals Inc. assisted in reviewing the manuscript. The study was supported by Regeneron Pharmaceuticals, Inc. and F. Hoffmann-La Roche Ltd.

## Notes

### Competing Interest Statement

ICMJE disclosure forms provided by the authors are available with the full text of this article.

### Clinical Trial

NCT04666441

### Author Declarations

Ethics approval was obtained from the following ethics review boards: Western Institutional Review Board, Puyallup, WA, USA; Research Compliance Office, Stanford University, Palo Alto, CA, USA; Providence St. Joseph Health Institutional Review Board, Renton, WA, USA; and University of Maryland Baltimore Human Research Protection Office, Baltimore, MD, USA.

