## Supplemental Files for "Phase 2 dose-ranging study of the virologic efficacy and safety of the combination COVID-19 antibodies casirivimab and imdevimab in the outpatient setting"

### Supplementary Appendix

1. Study sites and investigators
2. Regeneron study team
3. Quantitative virology assay
4. SARS-CoV-2 serology testing
5. Pharmacokinetic analysis methods
6. Additional statistical analysis methods
7. Study oversight
8. Supplementary figures

#### Appendix 1: Study sites and investigators

**AGA Clinical Trials, Miami, FL:** Dario Altamirano, Dickson Ellington

**APD Clinical Research, Splendora, TX:** Najmuddin Karimjee, Munib Daudjee, Jessica Laabs, Karen Marler, Natalee Calais, Zachary Sheppard, Kristy Carroll

**Arizona Liver Health, Tucson, AZ:** Anita Kohli, Vicki McIntyre, Yessica Sachdeva, Ashley Carney

**Arizona Liver Health, Mesa, AZ:** Yessica Sachdeva, Anita Kohli, Amanda McFarland, Dina Gibson, Victorine Ekoko

**Ark Clinical Research, Long Beach, CA:** Kenneth Kim, Jason Ahn, Nayna Paryani, Amber Mottola, Eva Day, Martha Navarro, Apinya Vutikullird

**Bio-Medical Research, LLC, Miami, FL:** Lilia Roque-Guerrero, Ana Gomez Ramirez, Javier Capote, Gisel Paz

**Carolina Medical Research, Clinton, SC:** Nancy Patel, Ravikumar Patel, Ryan Sattar

**Centex Studies, Lake Charles, LA:** Michael Seep, Celeste Brown, Joshua Whatley

**Chicago Clinical Research Institute, Downers Grove, IL:** Dennis Levinson, Azazuddin Ahmed, Norman James, Saad Alvi, Ann Kuehl

**Clinical Research of Central Florida, Winter Haven, FL:** Robinson Koilpillai, Stephanie Cassady, Jennifer Cox, Eduardo Torres

**Crossroads Clinical Research, Corpus Christi, TX:** Michael Winnie, Omesh Verma, Richard Leggett

**DM Clinical Research/BFHC Research, San Antonio, TX:** Ramon Reyes, Keith Beck, Brian Poliquin

**DM Clinical Research/LinQ Research, LLC, Pearland, TX:** Murtaza Mussaji, Jignesh Shah

**Duke Clinical Research, Durham, NC:** John Eppensteiner, Alexander Limkakeng, Samuel Francis

**Epic Medical Research, Red Oak, TX:** Haresh Boghara, Sunny Patel, Bari Eichelbaum

**Excel Clinical Research, Las Vegas, NV:** Duane Anderson, Sean Su, Alexander Akhavan, Joy Venglik, Diana Kirby, Crista Fedora

**Florida Pulmonary Research Institute, LLC, Winter Park, FL:** Faisal A. Fakih, Faisal M. Fakih, Fernando Alvarado, Daniel Layish, Jose Diaz, Andres Perez

**Future Innovative Treatments, LLC, Colorado Springs, CO:** Bhaktasharan Patel, Gary Tarshis

**Global Clinical Professionals Research, St Petersburg, FL:** Roxana Stoici, Gualberto Perez, Joseph Pica, Enrique Villareal

**Harlem Hospital Center – NYCHHC, Harlem, NY:** Farbod Raiszadeh, Sharon Mannheimer, Khaing Myint, Lovelyamma Varghese, Hussein Assallum, Akari Kyawa, Karina Chan

**Holy Name Medical Center, Teaneck, NJ:** Suraj Saggar, Thomas Birch, Benjamin De La Rosa, Karyna Neyra, Erina Kunwar

**Hope Clinical Research, Canoga Park, CA:** Hessam Aazami, Cheryl Bland, Mary Michelle Nolasco

**Houston Methodist Hospital, Houston, TX:** Howard Huang, Jihad Georges Youssef, Simon Yau, Ahmad Goodarzi, Mukhtar Al-Saadi, Faisal Zahiruddin

**IACT Health, Columbus, GA:** Jeffrey Kingsley, April Pixler

**Innova Health Care Services (INOVA Fairfax Hospital), Falls Church, VA:** Christopher deFilippi, Christopher King, Lindsay Clevenger

**Maryland School of Medicine, Baltimore, MD:** Richard Wilkerson, Shivakumar Narayanan, Joel Chua, Jennifer Husson, John Baddley

**Mercury Clinical Research, Houston, TX:** Rajasekaran Annamalai, Huy Nguyen, Nizar Nayani, Mahalakshmi Ramchandra

**Miami Valley Hospital, Dayton, OH:** Thomas Herchline, Steve Burdette

**Midland Florida Clinical Research Center, Deland, FL:** Godson Oguchi, Judepatricks Onyema

**Midway Immunology and Research Center, Fort Pierce, FL:** Moti Ramgopal, Brenda Jacobs

**Next Level Urgent Care, Houston, TX:** Terence Chang, Robbyn Traylor, Lenee Gordon, John McDivitt, Lizette Castro

**Peninsula Research Associates, Rolling Hills Estates, CA:** Lawrence Sher, Monica Saad, LeighAnn Schmidt

**Pharma Tex Research, LLC, Amarillo, TX:** David Brabham, Tarek Naguib, Mark Sigler

**PMG Research of McFarland Clinic, Ames, IA:** Jennifer Killion, Rupal Amin, Timothy Lowry

**PMG Research of Wilmington, Wilmington, NC:** Kevin Cannon, Mesha Chadwick

**Providence Saint John’s Health Center, Santa Monica, CA:** Trevan Fischer, Terese Hammond, Anmol Rangoola

**Qway Research, Hialeah, FL:** Oscar Galvez, Fausto Castillo

**Remington-Davis, Columbus, OH:** Edward Cordasco, Brian Zeno, Heather Lee

**Ruane Clinical Research Group, Los Angeles, CA:** Peter Ruane, Peter Wolfe, Kenny Trinidad, Isaac Berlin

**San Francisco Research Institute, San Francisco, CA:** Mark Savant, Edna Yee

**SignatureCare Emergency Center – TC Jester, Houston, TX:** Alan Skolnick, Harold Minkowitz, David Leiman

**Stanford University, Palo Alto, CA:** Upinder Singh, Yvonne Maldonado, Jason Andrews, Chaitan Khosla, Hector Bonilla

**Tandem Clinical Research, Maitland, FL:** Esteban Olivera, Mayra Abreu

**Tandem Clinical Research, Marrero, LA:** Adil Fatakia, Marissa Miller, Kristen Clinton, Gary Reiss

**Triple O Research Institute PA, West Palm Beach, FL:** Olayemi Osiyemi, Jose A. Menajovsky-Chaves, Christina Campbell, Stephanie Martinez

**Universal Medical and Research Center, LLC, Miami, FL:** Gerard Acloque**,** Agustin Martinez

**University of South Florida, Tampa, FL:** Kami Kim, Seetha Lakshmi, Asa Oxner, Jason Wilson, Lucy Guerra, Tiffany Vasey, Susannah Hall

**Willis-Knighton Physician Network, Shreveport, LA:** Joseph Bocchini, Clint Wilson

#### Appendix 2: Regeneron study team

Kathryn Adams, Dawn Applegate, Dhanalakshmi Barron, Mary Basilious, Alina Baum, Travis Bernardo, Eleonora Bianchi, Dona Bianco, Manika Bista, Mandi Blackmon, Teresa Blake, Jessica Boarder, Lisa Boersma, Denise Bonhomme, Peter Boutros, Derrick Bramble, Ned Braunstein, Aurora Breazna, Alison Brown, Elizabeth Bucknam, Cynthia Portal-Celhay, Tyrah Chatman, Ravikanth Chava, Donna Cohen, Nikki Covino, S. Balachandra Dass, Amy Davis, John D Davis, Jeanelle De Villiers, Sherrie DeGuzman, Monica DeYoung, Thomas DiCioccio, Marc Dickens, Lacey Douthat, Ajla Dupljak, Will Eagan, Adil Fatakia, Joseph Fitzgerald, Kyle Foster, Samit Ganguly, Paul Gao, Peter Gasparini, Evelyn Gasparino, Heath Gonzalez, Ruchin Gorawala, Lilia Roque-Guerrero, Daya Gulabani, Mary Hasinsky, Sheree Hairston, Jennifer D Hamilton, Robert Hamlin, Dawlat Hassan, Russell Haywood, Brian Head, Philippa Hearld, Ingeborg Heirman, Gary Herman, Olga Herrera, Andrea T Hooper, Romana Hosain, Susan Irvin, Ramya Iyer, Lisa Jackson, Rohit Kamath, Wendy Kampman, Helen Kang, Denise Kennedy, Yunji Kim, Elisa King, Michael Klingler, Bari Kowal, Mi Young Kwon, Christos A Kyratsous, Carol Lee, Maria Lichtschein, David Liu, Leah Lipsich, Kristy Macci, Adnan Mahmood, Marco Mancini, Nagaratna Reddy Medapti, Nilang Mehta, Colin McDonald, Kristina McGuire, Stephanie Miller, Nkechi Moghalu, Kosalai Mohan, Nicholas Moore, Bret J Musser, Emily Nanna, Eduardo Forleo-Neto, Thomas Norton, Soraya Nossoughi, Esther Huffman O'Keefe, Cynthia Pan, Carrie Papazian, Janie Parrino, Michael Partridge, Christina Perry, Cynthia Plante, Cynthia PortalCelhay, Kalpana Pullakhandam, Emina Radoncic, John Rembis, Nelson Rita, Mivianisse Rodriguez, Neena Sarkar, Viral Seth, Ileana Schirmer, Liyang Shao, Shelley Geila Shapiro, Matthew Silverman, Carmella Simiele, Isarael Simonetti, Lisa Sherpinsky, Anne Smith, Jasmine Smith, Pamela Snodgrass, Neil Stahl, Yuhwen Soo, Roxana Stoici, Michel Tarabocchia, Karen Thabet, Yanmei Tian, Caryn Trbovic, Ngan Trinh, Kenneth C Turner, Violet Vincent, Jeff Watts, Jianguo (Victor) Wei, David M Weinreich, Collen Whetzel, Joseph Wolken, George D Yancopoulos, Karen Yau, Ori Yellin, Jeannie Yo, Yuming Zhao, Bryan Zhu.

#### Appendix 3: Quantitative virology assay

Nasopharyngeal swabs were collected in 3 mL Viral Transport Medium (Catalog No. R99, Hardy Diagnostics, Santa Maria, CA, USA) at baseline (day 1) and study days 3, 5, 7, 15, and 22. Virologic testing for SARS-CoV-2 infection was performed at Viracor Eurofins (Lee’s Summit, MO, USA) by quantitative real-time RT-PCR (RT-qPCR). RNA was extracted from clinical samples with the Applied Biosystems™ MagMAX™ Viral/Pathogen II (MVP II) Nucleic Acid Isolation Kit (Catalog No. A48383, ThermoFisher, Waltham, MA, USA) on the ThermoFisher KingFisher Flex. Viral nucleic acids were detected and quantified using a SARS-CoV-2 RT-qPCR swab assay (Viracor Eurofins) which is performed using oligonucleotide primers and TaqMan™ probes for the detection of two regions of the viral N protein gene region of SARS-CoV-2 and an internal extraction and amplification control target on ABI 7500 SDS Instruments (Applied Biosystems, Waltham, MA, USA). Assay data are presented in units of copies (genomic equivalents) of SARS-CoV-2 per mL of Viral Transport Medium (VTM) yielded from the collection of nasopharyngeal swab specimens (copies/mL). The limit of detection was 299 copies/mL (2.47 log_10_ copies/mL), the lower limit of quantification was 714 copies/mL (2.85 log_10_ copies/mL), and the upper limit of quantification (ULoQ) was 7.1x10^7^ copies/mL (7.85 log_10_ copies/mL). Samples with results above the ULoQ were diluted and retested with the same assay. Analysis-positive RT-qPCR results (detected) below the lower limit of quantification were imputed as half of the lower limit of quantification (357 copies/mL) and negative RT-qPCR results (not detected) were imputed as 0 log_10_ copies/mL (1 copy/mL).

#### Appendix 4: SARS-CoV-2 serology testing

Serum was collected from patients at baseline. The EUROIMMUN Anti–SARS-CoV-2 ELISA (IgG) and the EUROIMMUN Anti–SARS-CoV-2 ELISA (IgA) assays were validated and run at ICON Laboratories (Farmingdale, NY) to detect endogenous anti-S1 protein antibodies. In addition, the Abbott Architect SARS-CoV-2 IgG assay was validated and run at ICON Laboratories to detect endogenous anti-nucleocapsid antibodies.

#### Appendix 5: Pharmacokinetic analysis methods

*Bioanalytical methods*

The concentrations of casirivimab (REGN10933) and imdevimab (REGN10987) in human serum were measured using validated immunoassays which employ streptavidin microplates from Meso Scale Discovery (MSD, Gaithersburg, MD, USA). The methods utilized two anti-idiotypic, monoclonal antibodies, each specific for either casirivimab or imdevimab as the capture antibodies. Captured total casirivimab and total imdevimab were detected using two different, non-competing, anti-idiotypic monoclonal antibodies, each also specific for either casirivimab or imdevimab. In these total assays, concentrations of total casirivimab or total imdevimab were measured without regard to binding site occupancy. The assays detect antibody with either 1 or 2 unoccupied binding sites, as well as antibody with both binding sites occupied. The bioanalytical methods specifically quantified the total concentrations of each anti–SARS-CoV-2 spike monoclonal antibody separately, with no interference from the other antibody. The assays have a lower limit of quantification (LLoQ) of 0.156 mg/L for each analyte in the undiluted serum sample.

*Pharmacokinetic methods*

The pharmacokinetics (PK) analysis population included all participants in the modified full analysis set (mFAS) who received active study drug and who had at least 1 non-missing total analyte (total casirivimab or total imdevimab) concentration result following dosing with study drug (modified concentration-response analysis set, or mCRAS). Participants were analysed based on actual treatment received. Overall, 429 patients were included in the PK analysis set.

Blood samples for measurement of casirivimab and imdevimab concentrations in serum were collected from patients at predose, within 60 min after the end of infusion (intravenous [IV] only) or at least 1 h after study drug administration (subcutaneous [SC] only) and on days 3, 5, and 7.

Concentrations of total REGEN-COV (sum of casirivimab + imdevimab) in serum were used for all PK and concentration-response analyses.

#### Appendix 6: Additional statistical analysis details

For between-group comparisons, the 95% CI half-width between any two treatment groups with this sample size would be 0.27 log_10_ copies/mL. The placebo IV and placebo SC arms were combined in the efficacy analyses of the viral load endpoints, as the route of administration does not alter the pharmacodynamic response of patients who received placebo. Active IV and SC REGEN-COV arms were not combined in any analysis, including for corresponding doses.

The efficacy analyses were based on observed data, with no imputation for missing data. Viral load values less than the LLoQ (714 copies/mL, equivalent to 2.85 log_10_ copies/mL) of the RT-qPCR assay, but with positive qualitative results, were set to half of the LLoQ of the RT-qPCR assay. Values with nondetectable RNA were set to 0 log_10_ copies/mL if the reason for the negative value was not a failed test. Viral load values above the upper limit of quantification were retested using the reflex test with diluted sample.

The primary efficacy variable was calculated using the linear trapezoidal rule to calculate area under the curve (AUC) and then time-weighted average daily change from baseline (TWACB) was determined using AUC. The calculation for time-weighted average (TWA) is the following:

$$TWA=\frac{AUC}{t_{k}-t_{1}}=[\sum_{i=2}^{k} \left( t_{i}-t_{i-1} \right)*(V_{i}+V_{i-1})/2]/(t_{k}-t_{1})$$

where 𝑉_𝑖_ is the result of change in viral load (log_10_ copies/mL) at time point 𝑡_𝑖_ from baseline. As baseline is time point t_1_, then 𝑉_1_ = 0.

The primary efficacy analysis was conducted using 803 randomised and treated patients enrolled in the study up to day 7, in order to evaluate approximately 400 patients who were seronegative at baseline. Active treatment arms were compared to the combined placebo group in descending order, as depicted below:

| Hierarchy Number | Description |
| --- | --- |
| 1 | 2400 mg IV versus pooled placebo |
| 2 | 1200 mg IV versus pooled placebo |
| 3 | 1200 mg SC versus pooled placebo |
| 4 | 600 mg IV versus pooled placebo |
| 5 | 600 mg SC versus pooled placebo |
| 6 | 300 mg IV versus pooled placebo |

#### Appendix 7: Study oversight

The trial was conducted in accordance with the principles of the Declaration of Helsinki, International Council for Harmonisation Good Clinical Practice guidelines, and applicable regulatory requirements. All patients provided written informed consent before participating in the trial. Ethics approval was obtained from the following ethics review boards: Western Institutional Review Board, Puyallup, WA, USA; Research Compliance Office, Stanford University, Palo Alto, CA, USA; Providence St. Joseph Health Institutional Review Board, Renton, WA, USA; and University of Maryland Baltimore Human Research Protection Office, Baltimore, MD, USA.

#### Appendix 8: Supplementary figures

##### Figure S1: Consort diagram of patient flow

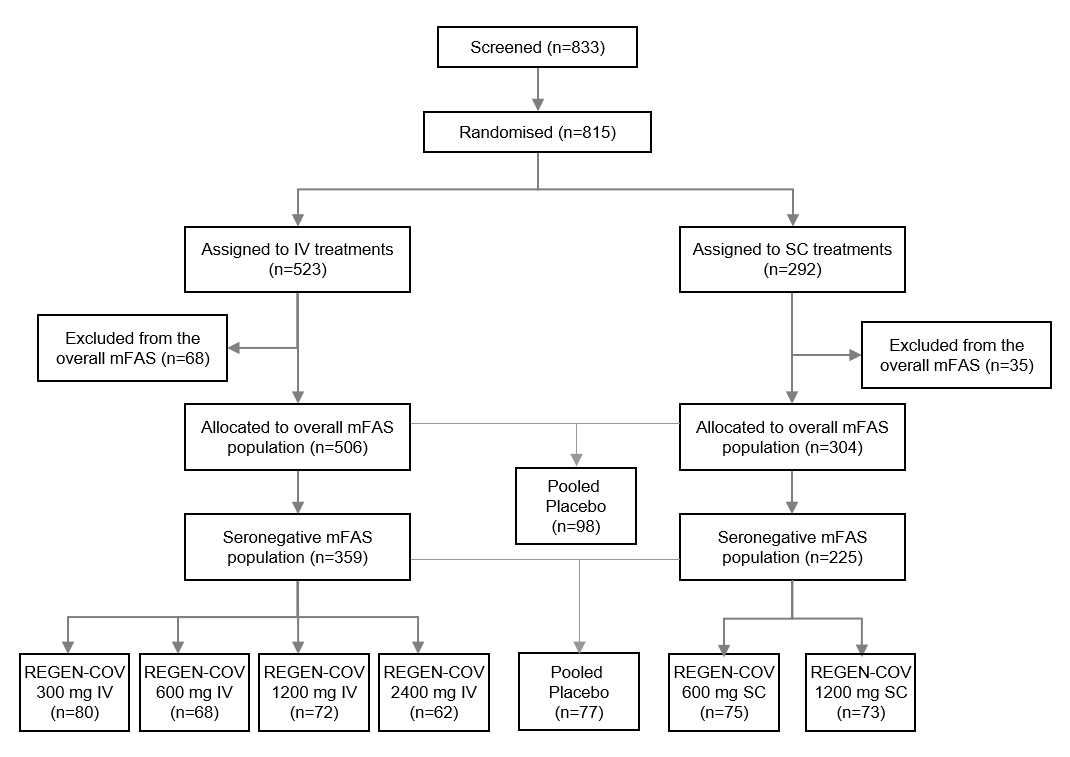

IV=intravenous; mFAS=modified full analysis set; SC=subcutaneous.
